# Quantifying social contact patterns in Minnesota during Stay-at-Home social distancing order

**DOI:** 10.1101/2021.07.12.21260216

**Authors:** Audrey M. Dorélien, Narmada Venkateswaran, Jiuchen Deng, Kelly Searle, Eva Enns, Shalini Kulasingam

## Abstract

SARS-CoV-2 is primarily transmitted through person-to-person contacts. It is important to collect information on age-specific contact patterns because SARS-CoV-2 susceptibility, transmission, and morbidity vary by age. To reduce risk of infection, social distancing measures have been implemented. Social contact data, which identify who has contact with whom especially by age and place are needed to identify high-risk groups and serve to inform the design of non-pharmaceutical interventions.

We estimated and used negative binomial regression to compare the number of daily contacts during the first wave (April-May 2020) of the Minnesota Social Contact Study, based on respondents age, gender, race/ethnicity, region, and other demographic characteristics. We used information on age and location of contacts to generate age-structured contact matrices. Finally, we compared the age-structured contact matrices during the stay-at-home order to pre-pandemic matrices.

During the state-wide stay-home order, the mean daily number of contacts was 5.6. We found significant variation in contacts by age, gender, race, and region. Adults between 40 and 50 years had the highest number of contacts. Respondents in Black households had 2.1 more contacts than respondent in White households, while respondents in Asian or Pacific Islander households had approximately the same number of contacts as respondent in White households. Respondents in Hispanic households had approximately two fewer contacts compared to White households. Most contacts were with other individuals in the same age group. Compared to the pre-pandemic period, the biggest declines occurred in contacts between children, and contacts between those over 60 with those below 60.

## INTRODUCTION

Prior to the availability of COVID-19 vaccines, public health officials and governments were reliant on non-pharmaceutical interventions (NPIs) to control and mitigate the spread of novel infections like SARS-CoV-2. School closures, physical distancing measures, and stay-at-home orders are all examples of NPIs that have been used to reduce the risk of transmission by limiting interpersonal contacts. Many infectious disease transmission models that seek to predict the disease trajectory, test the impact of various NPI control measures, and determine optimal vaccination strategies, include age-structured contact patterns as a key model parameter (1–4). This is because the force of infection is affected by heterogeneity in mixing patterns related to mixing within and between different age groups (5). It is also important to present age-structured patterns because COVID-19 morbidity and mortality patterns vary by age (6); and this allows us to identify age groups that are at high-risk for contracting and transmitting COVID-19 based on reported levels of contact. Unfortunately, the United States lacks data on interpersonal/social contact patterns. Very little data was collected prior to the onset of the current COVID-19 pandemic; the existing baseline data are either dated and focused on measuring the mean duration of contacts, focused on small populations, or do not provide detailed information for non-household settings (7–9). Researchers modeling SARS-CoV-2 transmission in the U.S. have relied on data from European countries or extrapolated estimates of what U.S. contact patterns may look like (10,11).

Our study makes many important contributions to the scientific literature on social contact patterns. We analyze data from the Minnesota Social Contact Study (MN SCS), which collected information on all age groups (children and adults) during the pandemic using a representative sample of the Minnesota non-institutional population. Other recent studies conducted in the U.S. focused only on adults (18 years and older) and did not use a population representative sample of the US or any state’s population (12). We identify structural (e.g., weekday versus weekend) and socio-demographic factors (e.g., race, region, etc.) that are associated with elevated number of contacts during the stay-at-home (SAH) order in Minnesota. We quantify the extent to which the SAH order altered the number and pattern of contacts by comparing the MN Wave 1 (SAH) matrix with pre-pandemic contact matrices. Specifically we compare our age-structured mixing patterns with mixing patterns from the United Kingdom (UK POLYMOD) in 2006 (13). The POLYMOD (Improving Public Health Policy in Europe through Modelling and Economic Evaluation of Interventions for the Control of Infectious Diseases) surveys are the most widely cited social contact surveys; they were conducted in 2006 in eight different European countries. We also compare our findings with a US synthetic contact matrix, based on data from POLYMOD surveys and data on US household composition, labor force participation, school enrollment rates, and population age structure (10). Finally we compare the contacts occurring in the home with a matrix generated from American Time Use Survey data (9). We highlight which age groups had the greatest changes in behavior pre and post SAH. Finally, we compare the MN SAH contact patterns and age-structured contact matrix with contact patterns from other movement control orders in other geographic settings.

## RESULTS

### Number of contacts

Figure 1 shows the histograms of reported contacts. The average number of daily contacts in the MN SCS Wave 1 sample is 5.6 (5.3 if using unweighted data), however the distribution of contacts is skewed (Skewness = 6.2 and Kurtosis = 51.8). The median number of daily contacts is 3.0; 82 percent of respondents reported six or fewer contacts (Figure 1 and Table 1). There is a long tail with 45 (2.2%) respondents reporting more than 30 contacts and a handful reporting more than 100 contacts.

**Table 1.**
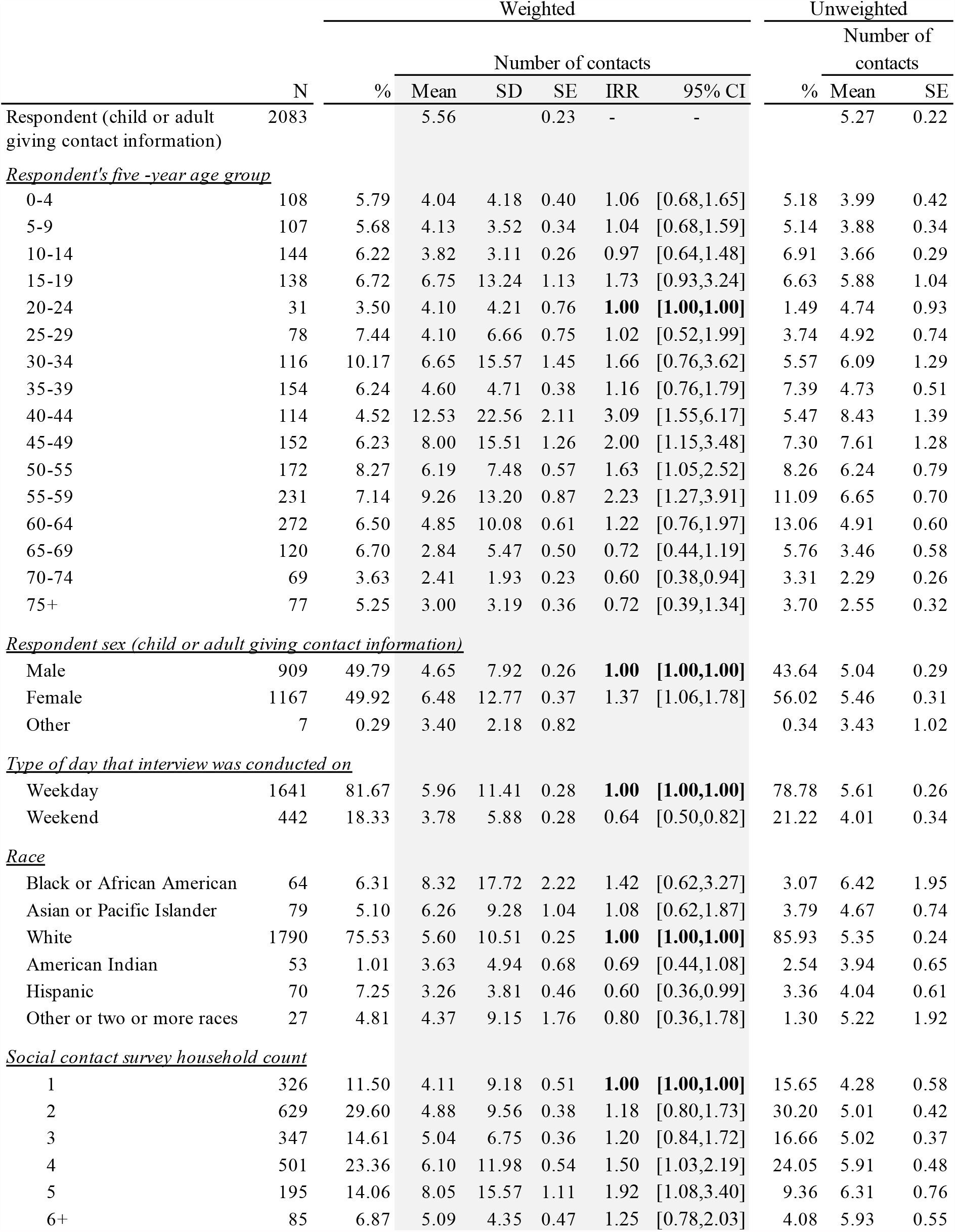
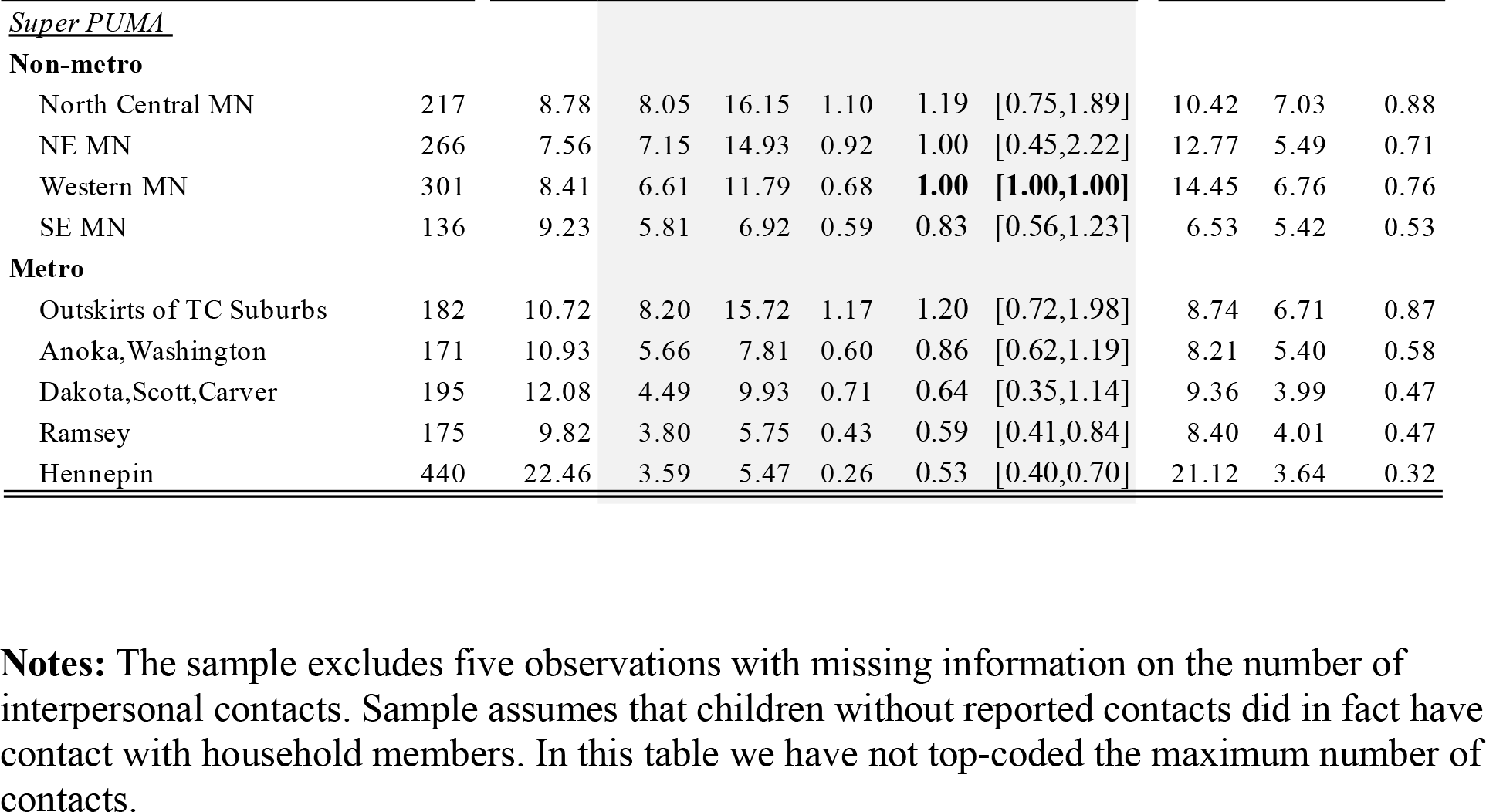
Descriptive statistics showing distribution of participants and daily contacts by demographic characteristics (N= 2,083).

**Figure 1.**
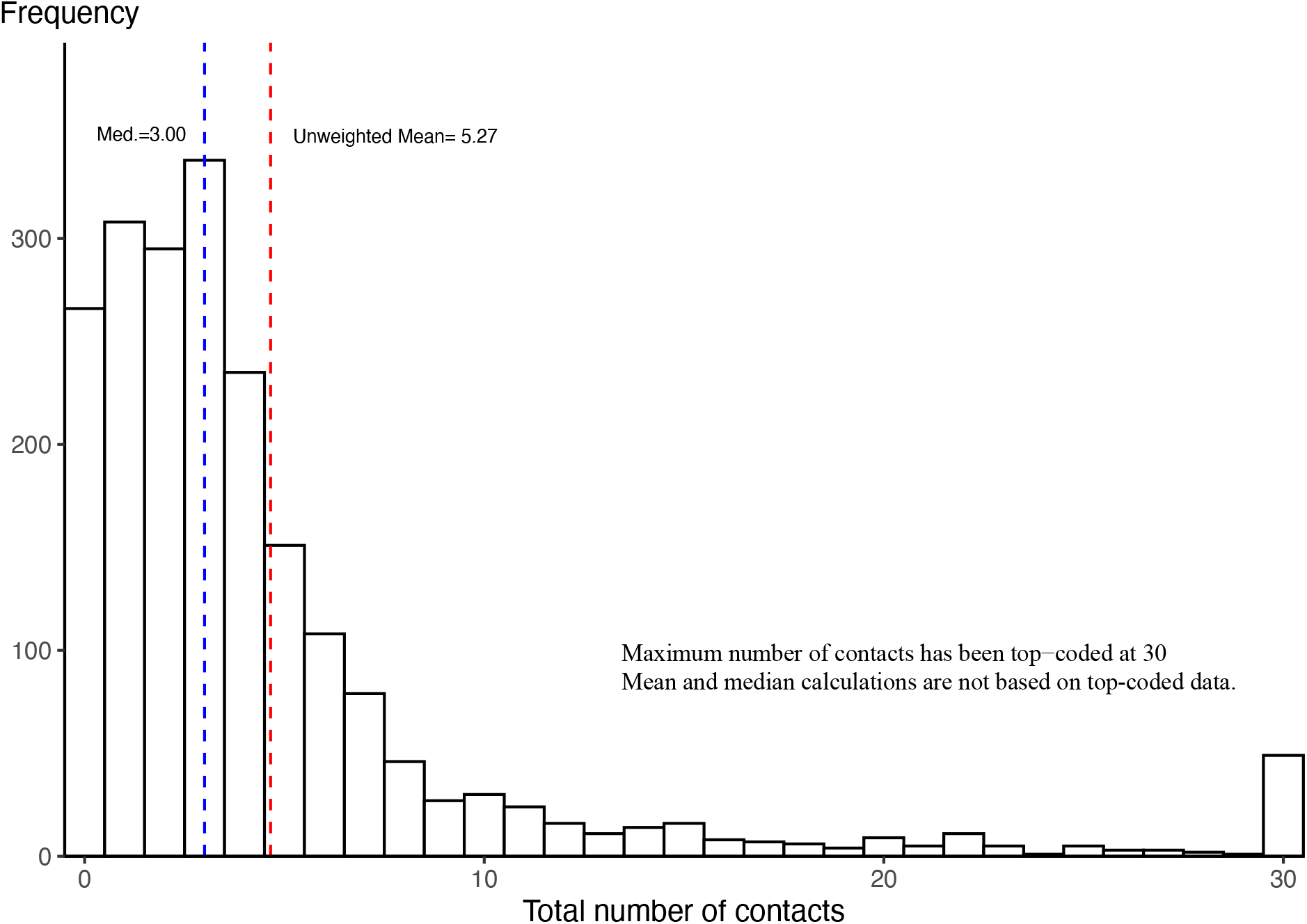
Histogram showing the distribution of the number of daily contacts.

### Demographic and structural factors associated with elevated contact rates

The 2,083 participants in our analytic sample recorded a total of 10,983 unique contacts. Frequency of social contacts were not homogeneous across age, gender, race, region, or type of day. Based on the weighted means in Table 1, older teens (15-19), and adults between the ages of 30-60 years had the highest number of daily contacts during the SAH order. On average, women had more daily contacts (6.5) compared to men (4.7) (IRR = 1.37; 95% CI = 1.06-1.78). Contacts were lower during the weekend compared to the weekday (IRR = 0.64; 95% CI = 0.50-0.82). Except for the suburbs of the twin cities (TC) of Minneapolis and Saint Paul, the average number of daily contacts was generally lower in metro areas compared to non-metro areas. An additional analysis shown in Appendix A reveals that for respondents in the metro areas the majority of contacts took place at home while for those in non-metro areas the majority of contacts took place at work and school. The daily number of contacts tended to increase as household size increased from one to four. We had relatively few large (household size >4) households therefore the confidence intervals were very large for these groups. Between April 17^th^ and May 17, 2020, in Minnesota, respondents in Black or African American households had approximately three (2.71) additional contacts (IRR = 1.42; 95% CI = 0.62-3.27) compared to respondents in White households. Respondents in Asian or Pacific Islander households had approximately the same number of contacts as respondent in White households (IRR = 1.08; 95% CI = 0.62-1.87). Respondents in Hispanic households had significantly fewer contacts compared to white households (IRR = 0.60; 95% CI = 0.36-0.99). The number of respondents in the “other or two or more race” category was extremely small; therefore, the estimates may not be reliable.

### Location of contacts

Figure 2 disaggregates the mean daily number of contacts by location (panel a) and displays the relative share (panel b) of contacts by location and respondent’s age group. For respondents younger than 25 and over 65, most contacts took place at the respondent’s home (or someone else’s home). Most contacts for respondents between the ages of 35-39 also took place in the home. This may be due to childcare duties during the SAH order. Children had a higher mean number of contacts taking place at home compared to adults; and adults aged 35-59 had a higher mean number of home contacts compared to adults 20-34 and 60 and older. The age pattern of home contacts was likely driven by differences in household composition over the life course. For working-age respondents (ages 15-69) approximately 30-66 percent of all contacts took place in the workplace. About 30% of all contacts for children under the age of five took place in daycare setting during the SAH order.

**Figure 2.**
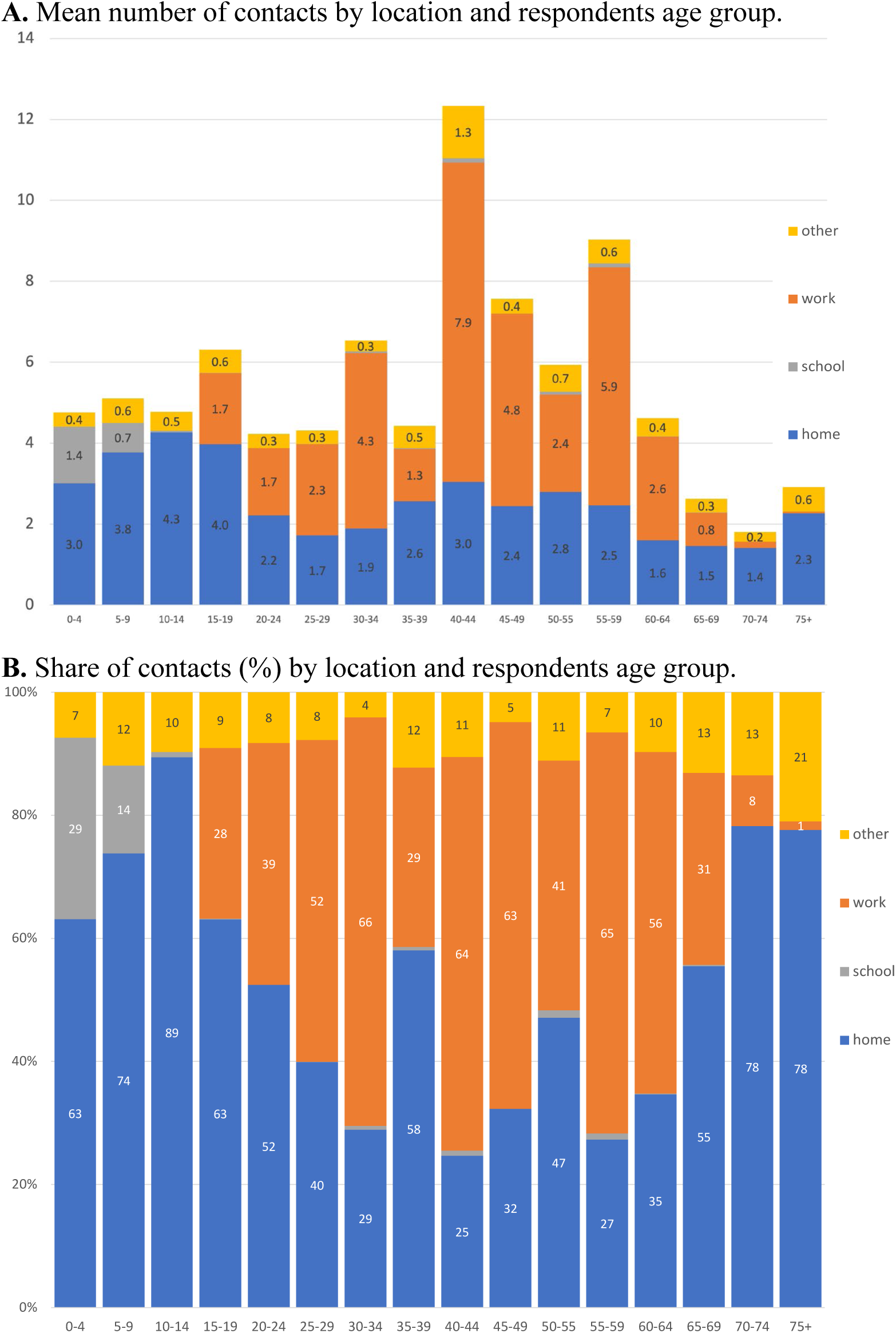
Age pattern of contacts by location, April 17-May 17, 2020. Note: For this analysis the maximum number of contacts for a respondent was not top coded at 30. Home category includes respondent’s own home and someone else’s home. The mean number of contacts in panel A may not equal the values in Table 1 because these were based on the detailed contact data and not the total number of reported contacts. Sample weights were used.

### Age-structured contact matrices results

To generate the age-structured contact matrices the maximum number of contacts that an individual could have was restricted to 30. This is in line with the Mossong (2008) POLYMOD matrices where the maximum number of contacts was limited to 29. Therefore, for these analyses the 2,049 participants had 9,618 unique contacts for which age was known or imputed.

Figure 3 presents the overall age-structured contact matrices as well as age-structured contact matrices for different locations. Overall, the age-assortative index *Q* of the SAH age-structured contact matrix measured in this study is equal to 0.21; in comparison, the *Q* index for the pre-pandemic UK POLYMOD matrix is 0.18. The greatest amount of mixing took place between respondents and contacts in the same age group. However, unlike in pre-pandemic matrices such as the UK POLYMOD matrix, there was not much variation in the level of age-assortative contacts for those below age 60 (range is between from 1.36 to 1.77 contacts). Children and young adults no longer had the largest number of assortative contacts (Appendix B1).

**Figure 3.**
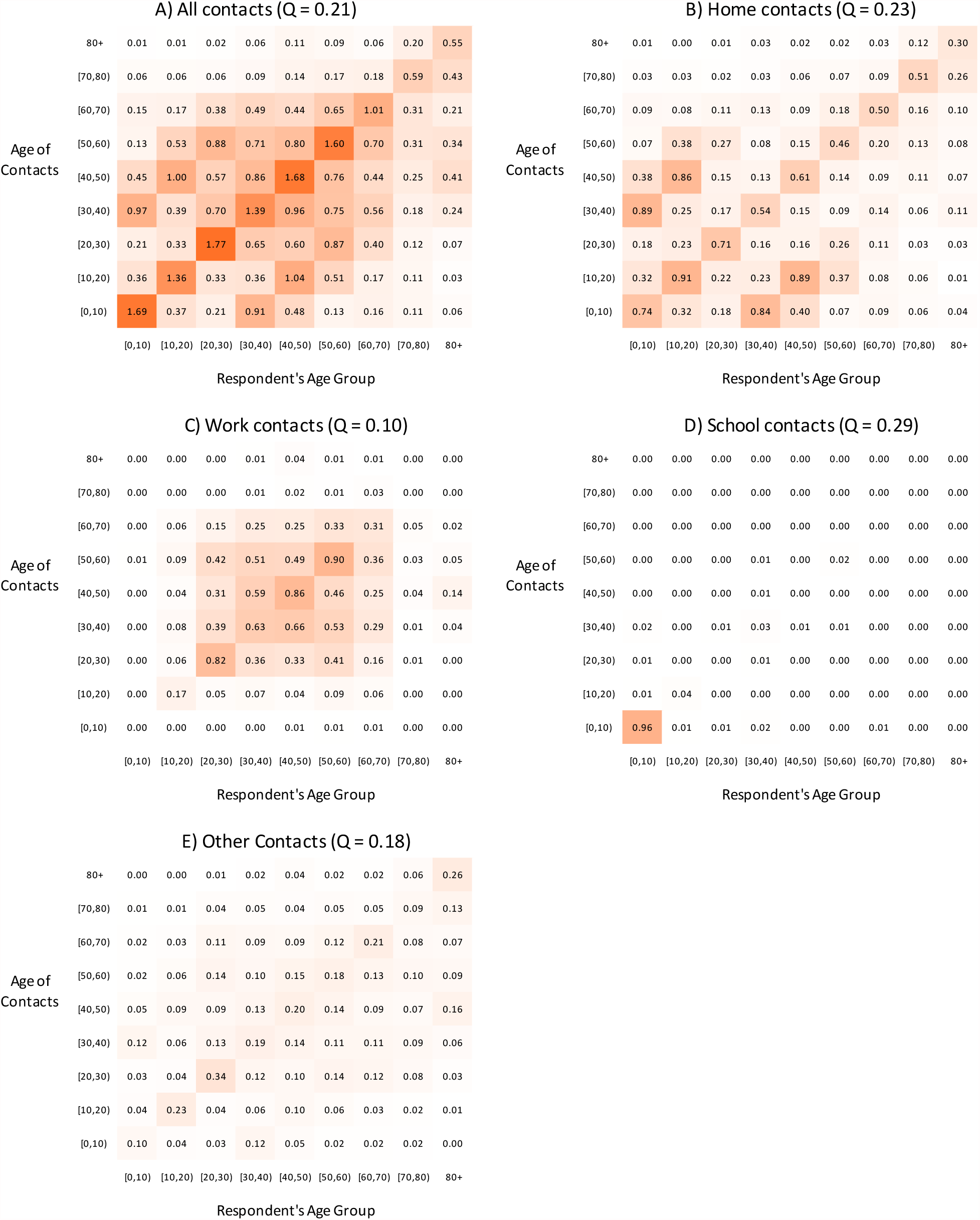
MN SCS Wave 1 (April-May 2020) age-structured contact matrix during SAH Order by location. Home contacts were defined as own-home contacts, and other contacts included transport, store, outdoors and other contacts not located at home, work, or school.

The SAH matrix also contain prominent parallel off diagonal elements starting in the 30-40 years age groups for both respondents and contacts. As in the pre-pandemic matrices, these represent intergenerational mixing, especially between young children and adults at home (Figure 3a and 3b).

### Age mixing patterns by location

The location-specific age-structured contact matrices corroborate the fact that during the SAH order most contacts took place at home (Figure 3b). The matrices and corresponding *Q* indices also illustrate that the pattern of contacts across the age groups varied by setting. Contacts at home were dominated by interactions between parents and children as well as interaction between people in the same age groups (Figure 3b). The nature of contacts also differed based on setting. Most physical contacts took place at home whereas contacts in other settings were primarily conversational (analysis not shown).

Work contacts were predominantly limited to individuals between 15 and 65 years old and were assortative by age (Figure 3c). During the SAH order, schools were closed; however, some daycare facilities remained opened because childcare providers were considered an essential service (14). Consequently, the “school” matrix shows that children below age five were mixing with other children (physical and conversational) and had contact with parents/guardians and childcare providers (Figure 3d). There were small number of contacts between adults, which most likely represent mixing between childcare providers and interactions between childcare providers and parents/guardians.

The “other” location matrix represents contacts that took place outside of home, school, or work. These contacts could take place while commuting, in stores, outdoors, and or in someone else’s home. The magnitude of contacts between people taking place in other locations is relatively low (Figure 3e). Although the age-assortative mixing are the most prominent mixing patterns in these settings, the difference when compared to interactions between people of different age groups was low (Q = 0.18). One of the most striking features of the “other” matrix was that people over 50 had almost no interactions with children under 10.

### Comparisons with pre-pandemic patterns: UK POLYMOD and US Home matrix

If we assume that MN pre-pandemic contact patterns were like those of the United Kingdom, as measured in the UK POLYMOD survey then as a result of the pandemic and SAH order, the mean number of contacts shrunk for the entire population (with one exception) but there was also a change in age-based mixing patterns, therefore the reduction was not the same across every cell (Appendix B1). The greatest change was a reduction in school children interacting with children their own age and interacting with young adults (67-83% reduction), reflecting the closure of schools during SAH. There was also a large reduction in contacts between non-institutionalized individuals aged 60+ years and those in younger age groups (on average a 70% reduction). The change in the mean number of contacts among adults between 20-60 years old was lower (on average 43% reduction). Many of these patterns are also evident if we compare the MN SAH matrix to the US pre-pandemic synthetic matrix from Prem, Cook, and Jit (2017) (Appendix B2). The main difference is that when we compare with the pre-pandemic US synthetic matrix, we see an increase in the number of contacts between working age respondents and those above 65; this is an artefact because the synthetic matrix assumed no work contacts for those above 65.

On average, there was an increase in the number of contacts taking place at home. However, the number of contacts at home varied during the SAH depending on respondent’s age group. The number of contacts with others in the same age group increased (Appendix B3, see cells with black border). The number of home contacts between 65–75 years-old respondents and children 0-15 years-old also increased; this could represent grandparents stepping in to provide childcare during the stay-at-home order, and combining of households to make this possible. The number of interactions between adults and children also increased, with the exceptions of interactions with children 0-5 years old, which decreased. Overall, there appears to be a decline in the number of contacts between respondents younger than 55 and children between the ages of 0-5 years. Some of the declines in number of contacts taking place in the home may be due to data quality issues. There is evidence that the MN SCS Wave 1 survey may underestimate the number of household contacts. As mentioned in the materials and methods section, there were children for which parents stated that they had zero contacts; in the data cleaning process we assumed that these children had contacts with household members. Additionally, there were still 85 adults who did not live alone yet reported zero contacts with household members.

## DISCUSSION

We conducted one of the first social contact survey in the United States that includes children and includes a population that is representative at a state level. The age-structured contact matrices are a key input for modeling COVID-19 transmission dynamics and may also be useful for other respiratory infectious diseases such as influenza and pertussis.

During the stay-at-home orders implemented during the first wave of the COVID-19 pandemic, the mean number of contacts in MN was 5.6 (median = 3). Our results are in line with wave 1 of the Berkeley Interpersonal Contact Survey (BICS), a non-probability sample of adults 18+ in 6 US cities conducted from April 10^th^ to May 4^th^ (Feehan and Mahmud, 2020) which also reported a median of three daily contacts. If we assume that the pre-pandemic mean daily number of contacts was around 13, the POLYMOD countries average (Mossong et al. 2008), then the lockdown significantly reduced the number of daily contacts. However, the MN mean daily number of contacts during the SAH order was higher than those in some other countries during their respective orders/lockdowns, which were more restrictive. For instance, the mean daily contacts during the United Kingdom’s lockdown was 2.8 daily contacts (Jarvis et al. 2020); 3.2 in Luxembourg (15); and 2.0 in Wuhan, China (16).

It is important to identify demographic groups that have high contact rates. Our results show that the mean number of social contacts during the SAH order were not homogeneous across region, race, gender, and age. We find some spatial differences in the mean number of daily contacts. During the first wave of the pandemic and SAH, on average respondents in non-metro areas had more contacts than those in the metro areas, and most of these contacts were at work or school. These spatial differences could be due to differences in occupation and respondent adherence to SAH orders. Another explanation is that during this period, COVID-19 cases were not as prevalent in non-metro settings (17).

The study found greater interpersonal contacts for Black households compared to White households, from which we can infer a greater likelihood of COVID-19 exposure. Exposure and rates of infections are of course also driven by the type of work, multigenerational households, and other factors. Nevertheless, although we found a pattern, the result was not statistically significant. In addition, the variation in the number of contacts for respondents in Black households was large (Standard Deviation = 17.7) compared to other racial/ethnic groups (White Standard Deviation= 10.5). One explanation could be that Black respondents are more likely to be essential workers and engaged in work in the health/eldercare industry (18). In our survey, respondents from Hispanic households had fewer interpersonal contacts; this may be because they were more likely to work in restaurants and thus vulnerable to being laid off and likely to work in restaurants (18). A deeper exploration of racial and ethnic differences in contact patterns is needed, as is an assessment of differences by employment status, occupation, income, and household composition. Before adjusting for sampling weights, 86 percent of the respondents in the MN SCS sample were White households (Table 1). BIPOC communities in MN also have a different age distribution and are more likely located in metro areas. As such, future studies will need to oversample Black, Indigenous, and other communities of color accounting for these differences to obtain more robust estimates of contacts in these communities.

The magnitude of contacts and the mixing patterns are not homogeneous by age. The regression analysis and age-structured matrices, both highlight the central role of individuals in the 40–45-year age group. Individuals in this age group had the highest mean number of contacts (weighted mean = 12.69); this is primarily driven by the presence of seven outliers with more than 75 work contacts. Nevertheless, this age group also had a relatively large number of contacts at home and in other locations (Figure 2a). Individuals 40-50 years old have many age-assortative contacts in addition to contacts with people who are slightly older and younger (Figure 3a). They also have a lot of contacts with individuals between the ages 5-20 who are presumably their children (Table 1, Figure 3). Importantly, this group appears to have had the smallest reduction in contacts during the SAH order (Appendix B1 and B2), which suggests that future interventions that are based on social contact patterns should focus on this group, assuming our findings are replicated by others and in future waves of this survey. Policy makers should also pay attention to individuals between 45-65 years (a.k.a. the squeezed/sandwich, generations who must take care of children and elderly parents) because those aged 70+ years and younger adults reported many contacts with this age group.

While it is important to identify age groups that have high contact rates, it is also important to identify groups that may be the main drivers of disease transmission, since these may not always be the same. Disease transmission depends on the age-pattern of contacts and the age-pattern of susceptibility and infectiousness. One way to do this is to incorporate age-structured contact matrices in **models of infectious disease transmission**. The age-structured contact matrices generated in this paper have been used by the MN COVID-19 modeling team, a collaboration between the University of Minnesota School of Public Health and the Minnesota Department of Health. Specifically it is possible to analyze how changes in contact rates impact the reproductive number (R0) of respiratory pathogens by comparing the dominant eigenvalues of the age-structured contact matrices (12). Consequently, it is important to continuously measure contact patterns as they change, especially as different restrictions are lifted and as vaccines are rolled out to different age groups.

Our study has some important limitations. It is difficult to obtain detailed accurate information on all interpersonal contacts for respondents with large numbers of contacts. We capped the number of contacts respondents had to recall detailed information for at 30. We did this to limit the possibility of missing information and to reduce the burden on the respondent. To address this limitation, we collected information on the total number of contacts in different settings and developed a strategy to impute the missing reported ages at school and work for respondents with large numbers of contacts. Another limitation might include self-selection into the sample; the respondents might be more compliant with executive orders. Nevertheless, the self-selection into the sample occurred before the start of the pandemic. There may be lack of standardization in defining “effective contacts”; we did not take into consideration masking during the first wave of data collection. We believe that our findings, despite this approach, are conservative for three reasons. First, there is some evidence that the survey may underestimate the number of household contacts; many respondents who lived with others reported having zero contacts. Social desirability bias could also be a factor due to respondents wanting to appear to comply with the SAH order. In this survey we asked people to recall their contacts, therefore respondents may forget to include some contacts. However this error is likely small because respondents were asked to recall contacts from the previous day and during the lockdown most respondents had very few contacts (19,20).

In conclusion the MN SCS is an ongoing study monitoring social contact patterns in Minnesota during the COVID pandemic. It is one of first surveys in the US to collect information on both children and adults. We have shown that during the SAH order there were large reductions in number of daily contacts; and that there are significant differences in contact patterns based on respondents’ age, sex, gender, race/ethnicity, and region. We have also generated age-structured contact matrices which have been used in the MN COVID-19 modeling effort.

## MATERIALS AND METHODS

### Ethics Statement

Participation in the MN SCS survey was voluntary, and all analysis was carried out on anonymized data. The study was approved by the University of Minnesota Institutional Review Board PRF (0706S10181).

### Survey population

Wave 1 of the Minnesota Social Contact Study (MN SCS) was collected between April 17^th^ and May 17^th^, 2020. During this period the Emergency Executive Order 20-20, which directed Minnesotans to stay at home, was extended and the stay-at-home order was still in effect (14,21). This order mandated that Minnesotans, with the exceptions of essential workers, stay at home and practice physical distancing when in public. Also, any worker who can work from home, including essential workers in the critical sectors, were required to do so. Essential workers were classified according to the Department of Health as those employed in childcare, social work or administration related to these positions; critical infrastructure; farming; food production, retail, or essential retail; and health care, elder-care and individual or family care (14). The definition of an essential worker was broad and encompassed more than 75% of the workforce (22). The data collection period took place during the start of the first wave of infections in Minnesota. During the data collection period, the test positivity rate for SARS-CoV-2 reached a maximum of 15%, COVID-19 hospitalizations reached 9.8 weekly admissions per 100,000 residents, and there were 396 deaths (23).

### MN SCS weighting

The MN SCS drew its sample from past participants in the 2019 Minnesota Health Access Survey (MNHA) who indicated their willingness to participate in a follow up survey. Half of this population was randomly selected for Wave 1 of the MN SCS. Because the MN SCS draws its sample from the MNHA and can be thought of as a longitudinal sub-sample, we use the MNHA base weights to model the MN SCS base weights. We applied response propensity weighting to adjust for survey nonresponse. Most notably, the Survey of Income and Program Participation and the Medical Expenditure Panel Survey use this method to adjust their weights for attrition between rounds. We used logistical regressions of respondents’ decisions to continue in the sample and a set of demographic variables from the MNHA as covariates to model for respondents’ propensity of attrition (24,25). These covariates are age of the MNHA target and respondent, sex of the MNHA target and respondent, relationship between the MNHA target and the respondent, race and ethnicity, educational level, marital status, home ownership, country of origin, employment status, income, household size, area of residence (i.e., Census’ Public Use Microdata Area), and internet access.

Using these adjusted base weights, we post-stratify the base weights to the estimated population counts, mainly obtained from the 2018 American Community Survey restricted for Minnesota residents only. As in the MNHA, we follow a first post-stratification step for prepaid cell phones before appending the Landline and Cell frames. Then, we post-stratify the RDD and ABS samples independently using a set of demographic variables.^1^ Finally, we use the effective sample size composite to append both frames in the MN SCS and obtain the final MN SCS weights. In Table 1, we present the unweighted and weighted distribution of the sample based on certain characteristics. The MN SCS Wave 1 over sampled non-metro regions. White and Native American households were overrepresented, while Black, Asian, and Hispanic households were under-represented.

### Survey Questions

We adopted a similar definition of contacts as that used in the POLYMOD and many other contact studies (13,19,26). A contact was defined as **1) either** a two-way conversation with three or more words in the **<u>physical presence</u>** of another person, **or** for children who are not yet speaking, a one-way conversation in the **<u>physical presence</u>** of another person; or **2) <u>physical skin-to-skin contact</u>** (for example a handshake, hug, kiss or contact sports). These contacts are further classified as conversational or physical.

A key feature of the survey is that data were collected for both children and adults. Respondents were defined as adults who self-reported their contacts or children under age 18 whose contacts were reported by a household adult. All households with children under 18 were asked first to provide social contact information for a randomly selected child, followed by a request for the adult to continue and provide social contact data for themselves; consequently, some households had two respondents.

For each respondent in our survey, we collected information on their age. We then asked them to record the total number of contacts (overall and in school and work settings) in the previous day. Then, for up to 30 of their interpersonal contacts,^2^ we collected information on the contact’s age, gender, the location of the contact (home, school, work, transportation, etc.), whether the contact was physical or not; the duration, and the frequency with which they were usually in contact with this person. For respondents with a large number (10 or more) of school or work contacts, we collected aggregate information for the contacts in those settings. The survey and survey methodology are described in greater detail in Dorélien et al. (2020).

### Data quality and sample selection

A random sample of 2,790 households was drawn from MNHA for Wave 1 of data collection. The response rate was 57 percent. Data was collected on 1,602 households however 15 were dropped during data cleaning (27). The final sample contained 1,613 households with 2,088 respondents. In our analysis, we excluded and additional five respondents that did not report (refused or don’t know) the total number of interpersonal contacts. The final sample consisted of 2,083 participants (adults = 1,594, children = 489).

For respondents with more than 10 interpersonal interactions at work (work outliers = 147 respondents), we did not collect detailed age information on their work contacts. Therefore, we impute the ages of those missing work contacts for work outliers. Because of top coding (an upward bound on total number of contacts), we impute the ages of missing work contacts until a respondent has 30 detailed contacts. For instance, if a respondent provided detailed information about 10 contacts, but had 27 work contacts, we imputed the ages of the first 20 work contacts. Since we knew the total number of missing work contacts, we generated the age of each missing work contact by sampling contact’s ages from reported work contacts of other respondents who went to work, were in the same five-year age group, and were of the same gender as the respondent. This assumes that people of the same age and gender interact with a similar mix of people in the workplace. The age distribution of people going to work during the SAH order was the same for the work outliers and those with fewer than 10 work contacts. Based on balance tests, the only variable that was predictive of being a work outlier was “female”. We assumed that people of the same age and gender interact with a similar mix of people in the workplace. As a sensitivity test, we also imputed missing work contacts using predictive mean matching (PMM) imputation. There were no statistically significant differences between the age groups of contacts generated by the two methods.

We also did not collect detailed school contact data for respondents (n=17) with 10 or more contacts at school. We assumed that these contacts were of the same age as the respondents. During the data cleaning stage, we noticed that some (n=76) of the child participants reported no contacts. We assumed that children without reported contacts did not contacts outside the household but did still have contact with household members.

## Data Analysis

In this paper, we pooled all contacts (physical and conversational) and did not restrict contacts based on duration, or frequency of reported contacts. We focused on describing and quantifying age patterns of social contacts to capture potential infectious disease transmission pathways between age groups and to provide critical input data for mathematical models of SARS-CoV-2 transmission (Mossong et al. 2008).

## Descriptive Analyses

Data cleaning and analysis were conducted using Stata version 16 (29) and the *socialmixr* R package (30). First, we used histograms to show the distribution of mean daily contacts during the SAH. In Table 1, we tabulated the mean number of interpersonal contacts per day by respondent’s age group, gender, type of day, region, household race^3^, and household size. We display both the weighted and unweighted mean daily contacts, as well as the standard errors. To display variation, we also include the standard deviation. Finally, in Table 1, we also include the incidence rate ratio (IRR) estimated from a negative binomial regression that includes covariates for structural factors (survey mode and type of day). We cluster the standard errors at the household level to account for multiple respondents from the same households. Our regression includes sampling weights (31).

To understand how location influences the age pattern of contacts, we disaggregate the mean number of daily contacts by location and calculate the relative distribution of mean contacts in home, school, work, and other locations by respondent’s age group.

### Age-structured contact matrices (who interacts with whom)

We generated overall age-structured contact matrices as well as age-structured matrices by location using the *socialmixr* R package (30). Data imputation was conducted before applying the *socialmixr* package because, by default, the package excludes contacts with missing or refused age information.

Respondents and their contacts were grouped into five or ten-year age groups and include individuals between the ages of 0 and 80; consequently, the raw contact matrix displays the mean number of daily contact (*M*_*ij*_) between respondents in age group i and their contacts in age group j. We also accounted for sampling weights, and weights for the type of day (a weight of 5 for weekday and 2 for weekend). We calculate *M*_*ij*_ as:

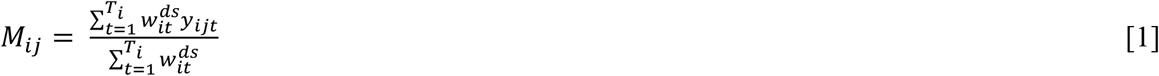

where *W*_*it*_ ^*ds*^ is the weights for the type of day and sampling weights combined for respondent t in age group *i, y*_*ijt*_ is the reported number of contacts made by respondent *t* in age group *i* with a contact in age group *j* and *T*_*i*_ represents all respondents in age group *i*.

Because of differences in reporting, interpersonal contacts in our dataset are not reciprocal. Therefore, we use population data from the 2019 MN American Community Survey to make sure that at the population level the matrices are symmetrical/reciprocal (32). This means that at a population level the total number of contacts made by respondents in age group *i* with contacts in age group *j* are the same as vice versa. We calculated the entries of the symmetric matrices 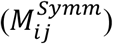 as:

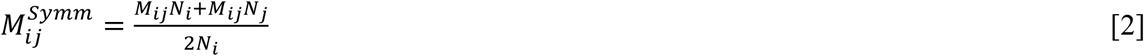

where *N*_*i*_ represents the sum of all individuals in age group *i* and age group *j* (2,30,32).

Finally for each age-structured contact matrix, we also calculated a measure of age-assortativeness using the index *Q*. If individuals interact solely with others in their age group, then the *Q* index takes a value of one; if there is homogeneous mixing the *Q* index takes a value of zero (33–35). The index is calculated by taking the trace of a matrix, *P*, whose elements represent the fraction of the **total contacts** of age group *i* with age group *j, P*_*ij*_ = *TT*_*ij*_/ ∑_*j*_ *T*_*ij*_. The matrix *T*_*ij*_ is obtained by multiplying 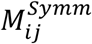 by a vector containing the number of people in each age group. The value of n represents the number of rows or columns of the *n* by *n* mixing matrix; in this study it is the number of age groups (Gupta, Anderson, and May 1989; Fava et al. 2021).

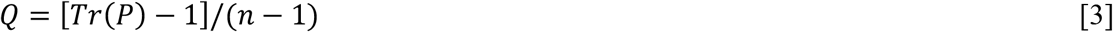

### Comparing matrices to published data and baselines (UK POLYMOD and ATUS home)

We compared our generated results to the UK POLYMOD matrices and calculated the percentage change in mean daily contacts. For our main analysis we focused on a comparison with the UK POLYMOD matrices in predicting pre-SAH contact rates. We also compared the home location matrix with the ATUS pre-pandemic home matrix, which is representative of the contiguous US states and is based on data from 2003-2018 for respondents aged 15 and older (9) and calculated the difference and percentage change in mean daily contacts. We hypothesized that the mean number of household/home contacts would increase during the pandemic. We also compare our results to the synthetic US matrices derived from Prem et al. (2017) and calculate the percentage change in mean daily contacts (Appendix B).

## Data Availability

The data is non-public and therefore cannot be shared at the individual level. We share the contact matrix data in the paper and upon request can share the corresponding confidence intervals that go with the estimated mean daily contacts between different age groups. The survey questionnaire is provided in the appendix of the Dorelien et al. 2020 paper.

https://doi.org/10.29115/SP-2020-0007

## Acknowledgements

We thank study participants. We also thank the following individuals at the Minnesota Department of Health: Alisha Simon, Sarah Hagge, Stefan Gildemeister, for assistance in creating the survey instrument and data cleaning. Giovann Alarcon Espinoza and Kathleen Call for generating sampling weights. Jason Kerwin for feedback on bootstrapping code. Participants in the 2021 Berkeley Formal Demography Workshop.

## Author contributions

**AMD:** survey instrument, conceptualization, supervising analysis, methodology, writing original draft, reviewing, and editing. **NV:** generated the age-contact matrices and histograms, assisted with writing original draft, reviewing and editing. **JD:** updating contact matrices, descriptive statistics, *Q* index, assisting missing value imputation, reviewing, and editing **KS:** survey instrument, PMM methodology, reviewing and editing writing. **EE:** funding acquisition, reviewing, and editing writing. **SK:** Funding acquisition, survey instrument, reviewing and editing writing

## Data availability

The data is non-public and therefore cannot be shared at the individual level. We share the contact matrix data in the paper and upon request can share the corresponding confidence intervals that go with the estimated mean daily contacts between different age groups. The survey questionnaire is provided in the appendix of the Dorélien et al. 2020 paper.

## Footnotes

1 These variables are age, sex, household size, home ownership, race and ethnicity at the household level (e.g., indicating the presence of racial and ethnic groups in the household), children’s presence in the household, highest educational level in the household, household size, internet access, and area of residence (i.e., Census’ Public Use Microdata Area).

2 The survey only collected detailed information on up to 30 contacts. Therefore, when we create the age-structured contact matrices, the number of contacts is top-coded at 30. It is common to top-code in these types of surveys. The POLYMOD surveys top-coded at 29 contacts (13,19).

3 Household race (HHRACE) is a variable created for the weighting process and is an indicator of whether anyone in the MNHA household is identified with a specific race or ethnic identity. Since members of a household may have multiple races or ethnic groups, HHRACE uses a hierarchy system to categorize these households: Hispanic, other race or multi-racial, American Indian, Asian or Pacific Islander, Black, and White. For example, if one member reports being Hispanic, and all other report a different ethnicity, the household is categorized as Hispanic. But if no one reports being Hispanic or multi-racial in the household, and one reports being Asian (non-Hispanic), the household is categorized as Asian or Pacific Islander.

## References

1. Jarvis CI, Van Zandvoort K, Gimma A, Prem K, Auzenbergs M, O’Reilly K, et al. Quantifying the impact of physical distance measures on the transmission of COVID-19 in the UK. BMC Medicine. 2020 May 7;18(1):124.

2. Feehan D, Mahmud A. Quantifying interpersonal contact in the United States during the spread of COVID-19: first results from the Berkeley Interpersonal Contact Study [Internet]. Epidemiology; 2020 Apr [cited 2020 Apr 23]. Available from: http://medrxiv.org/lookup/doi/10.1101/2020.04.13.20064014

3. Fava ED, Cimentada J, Perrotta D, Grow A, Rampazzo F, Gil-Clavel S, et al. The differential impact of physical distancing strategies on social contacts relevant for the spread of COVID-19. medRxiv. 2020 May 18;2020.05.15.20102657.

4. Medlock J, Galvani AP. Optimizing Influenza Vaccine Distribution. Science. 2009 Sep 25;325(5948):1705–8.

5. Del Valle SY, Hyman JM, Chitnis N. MATHEMATICAL MODELS OF CONTACT PATTERNS BETWEEN AGE GROUPS FOR PREDICTING THE SPREAD OF INFECTIOUS DISEASES. Math Biosci Eng. 2013;10(0):1475–97.

6. Boehmer TK, DeVies J, Caruso E, van Santen KL, Tang S, Black CL, et al. Changing Age Distribution of the COVID-19 Pandemic — United States, May–August 2020. MMWR Morb Mortal Wkly Rep. 2020 Oct 2;69(39):1404–9.

7. Zagheni E, Billari FC, Manfredi P, Melegaro A, Mossong J, Edmunds WJ. Using Time-Use Data to Parameterize Models for the Spread of Close-Contact Infectious Diseases. Am J Epidemiol. 2008 Nov 1;168(9):1082–90.

8. DeStefano F, Haber M, Currivan D, Farris T, Burrus B, Stone-Wiggins B, et al. Factors associated with social contacts in four communities during the 2007–2008 influenza season. Epidemiology & Infection. 2011 Aug;139(8):1181–90.

9. Dorelien A, Ramen A, Swanson I, Hill R. Analyzing the Demographic, Spatial, and Temporal Factors Influencing Social Contact Patterns in the U.S. and Implications for Infectious Disease Spread. Minnesota Population Center Working Paper Series [Internet]. 2020 [cited 2020 Nov 18]; Available from: https://assets.ipums.org/_files/mpc/wp2020-05.pdf

10. Prem K, Cook AR, Jit M. Projecting social contact matrices in 152 countries using contact surveys and demographic data. PLOS Computational Biology. 2017 Sep 12;13(9):e1005697.

11. Ewing A, Lee EC, Viboud C, Bansal S. Contact, Travel, and Transmission: The Impact of Winter Holidays on Influenza Dynamics in the United States. J Infect Dis. 2017 Mar 1;215(5):732–9.

12. Feehan DM, Mahmud AS. Quantifying population contact patterns in the United States during the COVID-19 pandemic. Nature Communications. 2021 Feb 9;12(1):893.

13. Mossong J, Hens N, Jit M, Beutels P, Auranen K, Mikolajczyk R, et al. Social Contacts and Mixing Patterns Relevant to the Spread of Infectious Diseases. PLOS Medicine. 2008 Mar 25;5(3):e74.

14. Walz T. Emergency Executive Order 20-48 Extending and Modifying Stay at Home Order, Continuing Temporary Closure of Bars, Restaurants, and Other Places of Public Accommodation, and Allowing Additional Workers in Certain Non Critical Sectors to Return to Safe Workplaces. Apr 30, 2020.

15. Latsuzbaia A, Herold M, Bertemes J-P, Mossong J. Evolving social contact patterns during the COVID-19 crisis in Luxembourg. PLOS ONE. 2020 Aug 6;15(8):e0237128.

16. Zhang J, Litvinova M, Liang Y, Wang Y, Wang W, Zhao S, et al. Changes in contact patterns shape the dynamics of the COVID-19 outbreak in China. Science [Internet]. 2020 Apr 29 [cited 2020 Jun 9]; Available from: https://science.sciencemag.org/content/early/2020/05/04/science.abb8001

17. Minnesota Department of Health. Weekly COVID-19 Report: 5/14/2020. 2020 May 14;15.

18. Minnesota Department of Health. Data by Race/Ethnicity [Internet]. COVID-19 Updates and Information - State of Minnesota. 2021b [cited 2021 May 25]. Available from: https://mn.gov/covid19/data/data-by-race-ethnicity/index.jsp

19. Hoang T, Coletti P, Melegaro A, Wallinga J, Grijalva CG, Edmunds JW, et al. A Systematic Review of Social Contact Surveys to Inform Transmission Models of Close-contact Infections: Epidemiology. 2019 Sep;30(5):723–36.

20. Coletti P, Wambua J, Gimma A, Willem L, Vercruysse S, Vanhoutte B, et al. CoMix: comparing mixing patterns in the Belgian population during and after lockdown. Scientific Reports. 2020 Dec 14;10(1):21885.

21. Walz T. Emergency Executive Order 20-20 Directing Minnesotans to Stay at Home. Mar 25, 2020.

22. Orenstein W. Who and what is and isn’t covered by Minnesota’s new ’stay-at-home’ order. MinnPost [Internet]. 2020 Mar 25 [cited 2021 May 25]; Available from: https://www.minnpost.com/state-government/2020/03/who-and-what-is-and-isnt-covered-by-minnesotas-new-stay-at-home-order/

23. Minnesota Department of Health. Public Health Risk Measures [Internet]. COVID-19 Updates and Information - State of Minnesota. 2021a [cited 2021 May 25]. Available from: https://mn.gov/covid19/data/response-prep/public-health-risk-measures.jsp

24. Weuve J, Tchetgen Tchetgen EJ, Glymour MM, Beck TL, Aggarwal NT, Wilson RS, et al. Accounting for bias due to selective attrition: The example of smoking and cognitive decline. Epidemiology. 2012 Jan;23(1):119–28.

25. Cole SR, Hernán MA. Constructing Inverse Probability Weights for Marginal Structural Models. Am J Epidemiol. 2008 Sep 15;168(6):656–64.

26. Leung K, Jit M, Lau EHY, Wu JT. Social contact patterns relevant to the spread of respiratory infectious diseases in Hong Kong. Scientific Reports. 2017 Aug 11;7(1):7974.

27. Dorélien AM, Simon A, Hagge S, Call KT, Enns E, Kulasingam S. Minnesota Social Contacts and Mixing Patterns Survey with Implications for Modelling of Infectious Disease Transmission and Control. Survey Practice. 2020 Aug 6;13(1):13669.

28. Davies NG, Klepac P, Liu Y, Prem K, Jit M, Eggo RM. Age-dependent effects in the transmission and control of COVID-19 epidemics. Nature Medicine. 2020 Jun 16;1–7.

29. StataCorp. Stata Statistical Software: Release 16. College Station, TX: StataCorp LLC; 2019.

30. Funk S. socialmixr: Social Mixing Matrices for Infectious Disease Modelling [Internet]. 2020 [cited 2020 Aug 3]. Available from: https://CRAN.R-project.org/package=socialmixr

31. Solon G, Haider SJ, Wooldridge JM. What Are We Weighting For? J Human Resources. 2015 Mar 31;50(2):301–16.

32. Klepac P, Kucharski AJ, Conlan AJ, Kissler S, Tang M, Fry H, et al. Contacts in context: large-scale setting-specific social mixing matrices from the BBC Pandemic project [Internet]. Epidemiology; 2020 Feb [cited 2020 Apr 6]. Available from: http://medrxiv.org/lookup/doi/10.1101/2020.02.16.20023754

33. Gupta S, Anderson R, May R. Networks of sexual contacts: implications for the pattern of spread of HIV. AIDS. 1989 Dec 1;3(12):807–17.

34. Iozzi F, Trusiano F, Chinazzi M, Billari FC, Zagheni E, Merler S, et al. Little Italy: An Agent-Based Approach to the Estimation of Contact Patterns-Fitting Predicted Matrices to Serological Data. PLOS Computational Biology. 2010 Dec 2;6(12):e1001021.

35. Fava ED, Adema I, Kiti MC, Poletti P, Merler S, Nokes DJ, et al. Individual’s daily behaviour and intergenerational mixing in different social contexts of Kenya. medRxiv. 2021 Mar 12;2021.03.10.21253281.

