## Supplementary figures and images for "Quantifying social contact patterns in Minnesota during Stay-at-Home social distancing order"

### Appendix A

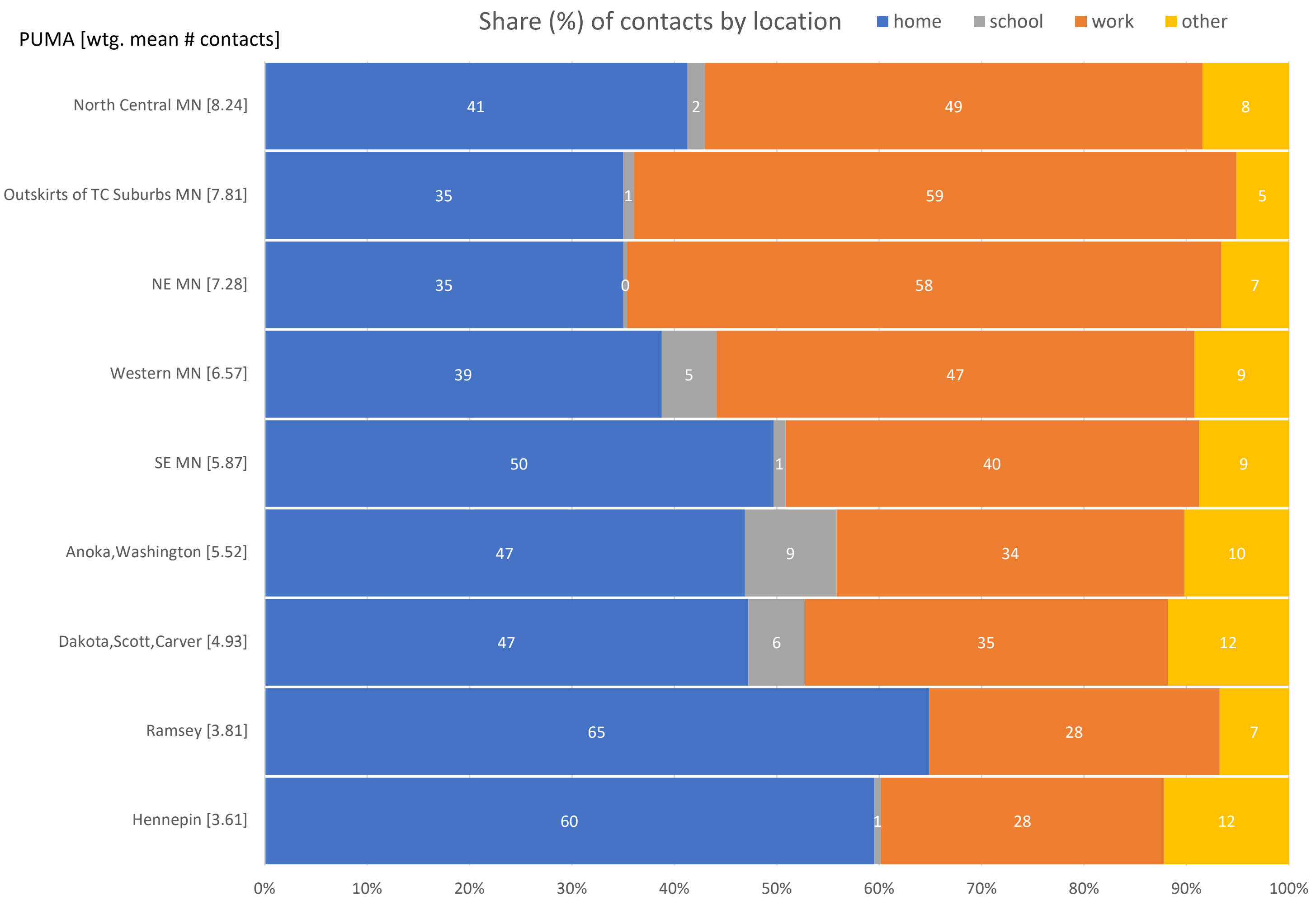
