## Appendix B1 for "Quantifying social contact patterns in Minnesota during Stay-at-Home social distancing order"

**Appendix B1.** Comparing UK POLYMOD and MN SCS Wave 1 contact matrices. In the percentage change matrix, if there was an increase in number of contacts during the SAH order, that value is coded in blue, if there was a decrease that value is in red.

**UK POLYMOD (Q = 0.18)**

Age of Contacts (rows) × Respondent's Age Group (columns)

| Age of Contacts | [0,10) | [10,20) | [20,30) | [30,40) | [40,50) | [50,60) | [60,70) | [70,80) |
|---|---|---|---|---|---|---|---|---|
| 80+ | 0.02 | 0.02 | 0.05 | 0.05 | 0.20 | 0.29 | 0.20 | 0.24 |
| [70,80) | 0.12 | 0.13 | 0.15 | 0.24 | 0.40 | 0.34 | 0.34 | 1.10 |
| [60,70) | 0.36 | 0.37 | 0.47 | 0.62 | 0.68 | 0.82 | 1.17 | 1.21 |
| [50,60) | 0.61 | 0.70 | 1.10 | 1.21 | 1.35 | 1.86 | 1.43 | 0.93 |
| [40,50) | 1.20 | 1.71 | 1.75 | 1.73 | 2.90 | 1.48 | 1.51 | 1.41 |
| [30,40) | 2.02 | 1.73 | 1.89 | 2.76 | 2.18 | 1.55 | 1.72 | 0.45 |
| [20,30) | 1.10 | 1.17 | 3.60 | 1.52 | 1.62 | 1.63 | 1.17 | 0.69 |
| [10,20) | 1.25 | 8.06 | 1.44 | 1.12 | 1.85 | 0.89 | 0.59 | 1.00 |
| [0,10) | 5.15 | 1.23 | 1.27 | 1.94 | 0.79 | 0.56 | 0.71 | 0.28 |

**MN W1 matrix (Q = 0.21)**

Age of Contacts (rows) × Respondent's Age Group (columns)

| Age of Contacts | [0,10) | [10,20) | [20,30) | [30,40) | [40,50) | [50,60) | [60,70) | [70,80) |
|---|---|---|---|---|---|---|---|---|
| 80+ | 0.01 | 0.01 | 0.02 | 0.06 | 0.11 | 0.09 | 0.06 | 0.20 |
| [70,80) | 0.06 | 0.06 | 0.06 | 0.09 | 0.14 | 0.17 | 0.18 | 0.59 |
| [60,70) | 0.15 | 0.17 | 0.38 | 0.49 | 0.44 | 0.64 | 1.01 | 0.32 |
| [50,60) | 0.13 | 0.53 | 0.88 | 0.71 | 0.80 | 1.60 | 0.69 | 0.31 |
| [40,50) | 0.45 | 1.00 | 0.57 | 0.87 | 1.67 | 0.76 | 0.44 | 0.25 |
| [30,40) | 0.97 | 0.39 | 0.70 | 1.37 | 0.98 | 0.75 | 0.56 | 0.18 |
| [20,30) | 0.21 | 0.33 | 1.75 | 0.66 | 0.60 | 0.87 | 0.40 | 0.12 |
| [10,20) | 0.36 | 1.36 | 0.33 | 0.36 | 1.04 | 0.51 | 0.17 | 0.11 |
| [0,10) | 1.69 | 0.37 | 0.21 | 0.91 | 0.48 | 0.13 | 0.17 | 0.11 |

**Percentage Change matrix**

Age of Contacts (rows) × Respondent's Age Group (columns)

| Age of Contacts | [0,10) | [10,20) | [20,30) | [30,40) | [40,50) | [50,60) | [60,70) | [70,80) |
|---|---|---|---|---|---|---|---|---|
| 80+ | 0.29 | 0.65 | 0.67 | -0.03 | 0.43 | 0.71 | 0.72 | 0.16 |
| [70,80) | 0.52 | 0.52 | 0.58 | 0.62 | 0.65 | 0.50 | 0.47 | 0.46 |
| [60,70) | 0.57 | 0.56 | 0.19 | 0.20 | 0.36 | 0.21 | 0.14 | 0.74 |
| [50,60) | 0.78 | 0.24 | 0.20 | 0.41 | 0.40 | 0.14 | 0.51 | 0.66 |
| [40,50) | 0.62 | 0.41 | 0.68 | 0.50 | 0.42 | 0.49 | 0.71 | 0.82 |
| [30,40) | 0.52 | 0.77 | 0.63 | 0.50 | 0.55 | 0.52 | 0.68 | 0.61 |
| [20,30) | 0.81 | 0.71 | 0.51 | 0.56 | 0.63 | 0.47 | 0.66 | 0.83 |
| [10,20) | 0.71 | 0.83 | 0.77 | 0.68 | 0.44 | 0.42 | 0.71 | 0.89 |
| [0,10) | 0.67 | 0.70 | 0.84 | 0.53 | 0.39 | 0.76 | 0.77 | 0.61 |

Fewer reduction in contacts for working-age adults (mean change = 43%)
