## Appendix B2 for "Quantifying social contact patterns in Minnesota during Stay-at-Home social distancing order"

**Appendix B2.** Comparing US Synthetic and MN SCS Wave 1 contact matrices. In the percentage change matrix, if there was an increase in number of contacts during the SAH order, that value is coded in blue, if there was a decrease that value is in red.

US Synthetic

From Prem et al. (2020)

| Age of Contacts | | | | | | | | | | | | | | | | |
|---|---|---|---|---|---|---|---|---|---|---|---|---|---|---|---|---|
| (75, 80) | 0.04 | 0.04 | 0.05 | 0.02 | 0.04 | 0.02 | 0.04 | 0.03 | 0.03 | 0.07 | 0.07 | 0.06 | 0.08 | 0.09 | 0.22 | 0.40 |
| (70, 75) | 0.07 | 0.05 | 0.06 | 0.03 | 0.05 | 0.03 | 0.04 | 0.07 | 0.07 | 0.07 | 0.08 | 0.10 | 0.19 | 0.23 | 0.68 | 0.25 |
| (65, 70) | 0.14 | 0.12 | 0.10 | 0.07 | 0.05 | 0.06 | 0.09 | 0.15 | 0.12 | 0.09 | 0.13 | 0.22 | 0.38 | 0.90 | 0.47 | 0.24 |
| (60, 65) | 0.17 | 0.17 | 0.10 | 0.10 | 0.12 | 0.16 | 0.22 | 0.25 | 0.21 | 0.18 | 0.31 | 0.55 | 1.07 | 0.48 | 0.48 | 0.17 |
| (55, 60) | 0.27 | 0.20 | 0.17 | 0.24 | 0.41 | 0.50 | 0.54 | 0.47 | 0.39 | 0.50 | 0.89 | 1.60 | 0.72 | 0.44 | 0.28 | 0.22 |
| (50, 55) | 0.34 | 0.24 | 0.32 | 0.53 | 0.74 | 0.91 | 0.80 | 0.86 | 1.04 | 1.08 | 1.93 | 1.22 | 0.55 | 0.35 | 0.33 | 0.38 |
| (45, 50) | 0.32 | 0.35 | 0.56 | 1.01 | 1.08 | 0.95 | 1.03 | 1.19 | 1.47 | 2.03 | 1.70 | 0.96 | 0.55 | 0.32 | 0.41 | 0.43 |
| (40, 45) | 0.50 | 0.85 | 1.05 | 1.13 | 0.92 | 1.13 | 1.32 | 1.83 | 2.52 | 1.42 | 1.52 | 1.09 | 0.63 | 0.54 | 0.55 | 0.34 |
| (35, 40) | 0.99 | 1.07 | 0.87 | 0.89 | 1.04 | 1.33 | 1.70 | 2.78 | 1.66 | 1.33 | 1.15 | 0.98 | 0.78 | 0.53 | 0.39 | 0.29 |
| (30, 35) | 1.06 | 0.89 | 0.54 | 0.64 | 1.15 | 1.73 | 2.72 | 1.57 | 1.45 | 1.15 | 1.16 | 1.23 | 0.72 | 0.50 | 0.23 | 0.23 |
| (25, 30) | 0.72 | 0.51 | 0.30 | 0.78 | 1.74 | 3.41 | 1.65 | 1.28 | 1.13 | 0.95 | 1.30 | 1.22 | 0.62 | 0.35 | 0.23 | 0.15 |
| (20, 25) | 0.41 | 0.19 | 0.35 | 1.60 | 4.26 | 2.02 | 0.99 | 0.71 | 0.87 | 0.89 | 1.02 | 0.78 | 0.42 | 0.28 | 0.15 | 0.14 |
| (15, 20) | 0.32 | 0.35 | 0.97 | 11.11 | 2.65 | 0.88 | 0.51 | 0.76 | 1.17 | 1.66 | 1.43 | 1.00 | 0.51 | 0.24 | 0.35 | 0.34 |
| (10, 15) | 0.50 | 1.22 | 8.28 | 3.07 | 0.33 | 0.21 | 0.71 | 0.91 | 1.01 | 0.75 | 1.04 | 0.75 | 0.34 | 0.36 | 0.32 | 0.42 |
| (5, 10) | 1.10 | 5.39 | 1.89 | 0.43 | 0.21 | 0.34 | 0.93 | 1.09 | 0.74 | 0.56 | 0.68 | 0.74 | 0.44 | 0.43 | 0.32 | 0.28 |
| (0, 5) | 2.60 | 0.99 | 0.30 | 0.17 | 0.26 | 0.58 | 0.69 | 0.67 | 0.34 | 0.37 | 0.31 | 0.54 | 0.45 | 0.27 | 0.11 | 0.21 |
| | (0, 5) | (5, 10) | (10, 15) | (15, 20) | (20, 25) | (25, 30) | (30, 35) | (35, 40) | (40, 45) | (45, 50) | (50, 55) | (55, 60) | (60, 65) | (65, 70) | (70, 75) | (75, 80) |

MN W1 matrix

Overall contacts with boostrap values

| | | | | | | | | | | | | | | | | |
|---|---|---|---|---|---|---|---|---|---|---|---|---|---|---|---|---|
| (75, 80) | 0.03 | 0.03 | 0.02 | 0.02 | 0.00 | 0.04 | 0.03 | 0.04 | 0.03 | 0.10 | 0.11 | 0.06 | 0.05 | 0.09 | 0.18 | 0.22 |
| (70, 75) | 0.02 | 0.04 | 0.04 | 0.04 | 0.04 | 0.04 | 0.07 | 0.04 | 0.07 | 0.09 | 0.08 | 0.09 | 0.09 | 0.16 | 0.50 | 0.26 |
| (65, 70) | 0.08 | 0.07 | 0.04 | 0.01 | 0.06 | 0.10 | 0.13 | 0.19 | 0.19 | 0.15 | 0.11 | 0.24 | 0.29 | 0.39 | 0.20 | 0.17 |
| (60, 65) | 0.09 | 0.07 | 0.07 | 0.18 | 0.22 | 0.27 | 0.34 | 0.26 | 0.35 | 0.23 | 0.43 | 0.45 | 0.83 | 0.36 | 0.15 | 0.12 |
| (55, 60) | 0.09 | 0.04 | 0.10 | 0.30 | 0.50 | 0.41 | 0.52 | 0.26 | 0.35 | 0.43 | 0.79 | 0.80 | 0.47 | 0.30 | 0.16 | 0.13 |
| (50, 55) | 0.08 | 0.06 | 0.15 | 0.54 | 0.39 | 0.47 | 0.34 | 0.32 | 0.45 | 0.45 | 0.88 | 0.70 | 0.39 | 0.12 | 0.12 | 0.23 |
| (45, 50) | 0.07 | 0.24 | 0.41 | 0.54 | 0.27 | 0.32 | 0.33 | 0.40 | 0.66 | 0.82 | 0.43 | 0.36 | 0.20 | 0.16 | 0.13 | 0.19 |
| (40, 45) | 0.21 | 0.42 | 0.61 | 0.45 | 0.33 | 0.31 | 0.34 | 0.65 | 1.19 | 0.73 | 0.47 | 0.32 | 0.33 | 0.23 | 0.11 | 0.07 |
| (35, 40) | 0.49 | 0.63 | 0.32 | 0.12 | 0.16 | 0.32 | 0.60 | 0.64 | 0.68 | 0.46 | 0.36 | 0.25 | 0.26 | 0.24 | 0.07 | 0.09 |
| (30, 35) | 0.56 | 0.25 | 0.11 | 0.19 | 0.35 | 0.53 | 1.00 | 0.61 | 0.37 | 0.39 | 0.38 | 0.52 | 0.35 | 0.17 | 0.11 | 0.08 |
| (25, 30) | 0.18 | 0.08 | 0.02 | 0.14 | 0.53 | 1.02 | 0.52 | 0.32 | 0.32 | 0.38 | 0.51 | 0.40 | 0.28 | 0.13 | 0.06 | 0.08 |
| (20, 25) | 0.09 | 0.06 | 0.14 | 0.45 | 1.17 | 0.48 | 0.30 | 0.14 | 0.32 | 0.28 | 0.39 | 0.45 | 0.20 | 0.07 | 0.06 | 0.01 |
| (15, 20) | 0.06 | 0.11 | 0.31 | 1.37 | 0.44 | 0.12 | 0.16 | 0.10 | 0.42 | 0.56 | 0.52 | 0.27 | 0.16 | 0.01 | 0.07 | 0.05 |
| (10, 15) | 0.14 | 0.41 | 0.81 | 0.34 | 0.15 | 0.02 | 0.11 | 0.31 | 0.63 | 0.47 | 0.16 | 0.09 | 0.07 | 0.05 | 0.06 | 0.04 |
| (5, 10) | 0.39 | 1.01 | 0.41 | 0.12 | 0.06 | 0.08 | 0.23 | 0.62 | 0.43 | 0.27 | 0.07 | 0.04 | 0.07 | 0.08 | 0.06 | 0.07 |
| (0, 5) | 1.61 | 0.38 | 0.13 | 0.06 | 0.09 | 0.17 | 0.51 | 0.45 | 0.20 | 0.07 | 0.08 | 0.08 | 0.09 | 0.09 | 0.04 | 0.06 |
| | (0, 5) | (5, 10) | (10, 15) | (15, 20) | (20, 25) | (25, 30) | (30, 35) | (35, 40) | (40, 45) | (45, 50) | (50, 55) | (55, 60) | (60, 65) | (65, 70) | (70, 75) | (75, 80) |

Respondent's Age Group

Percentage Change from US Synthetic to MNW1 matrix

(US Synthetic - MNW1)/US Synthetic

| | | | | | | | | | | | | | | | | |
|---|---|---|---|---|---|---|---|---|---|---|---|---|---|---|---|---|
| (75, 80) | 0.34 | 0.22 | 0.64 | -0.05 | 0.89 | -0.94 | 0.13 | -0.29 | 0.01 | -0.47 | -0.59 | 0.12 | 0.33 | 0.03 | 0.17 | 0.44 |
| (70, 75) | 0.68 | 0.27 | 0.39 | -0.35 | 0.16 | -0.36 | -0.66 | 0.42 | 0.07 | -0.28 | 0.00 | 0.01 | 0.52 | 0.33 | 0.28 | -0.02 |
| (65, 70) | 0.44 | 0.43 | 0.56 | 0.82 | -0.11 | -0.89 | -0.51 | -0.31 | -0.64 | -0.62 | 0.14 | -0.11 | 0.23 | 0.56 | 0.57 | 0.31 |
| (60, 65) | 0.47 | 0.58 | 0.32 | -0.91 | -0.81 | -0.68 | -0.58 | -0.02 | -0.62 | -0.27 | -0.39 | 0.17 | 0.23 | 0.26 | 0.69 | 0.30 |
| (55, 60) | 0.66 | 0.78 | 0.42 | -0.29 | -0.22 | 0.17 | 0.05 | 0.46 | 0.12 | 0.14 | 0.12 | 0.50 | 0.35 | 0.31 | 0.43 | 0.39 |
| (50, 55) | 0.76 | 0.74 | 0.53 | 0.00 | 0.47 | 0.49 | 0.58 | 0.63 | 0.57 | 0.58 | 0.54 | 0.43 | 0.29 | 0.65 | 0.64 | 0.41 |
| (45, 50) | 0.79 | 0.31 | 0.27 | 0.47 | 0.75 | 0.66 | 0.68 | 0.67 | 0.55 | 0.60 | 0.75 | 0.62 | 0.63 | 0.50 | 0.67 | 0.55 |
| (40, 45) | 0.59 | 0.51 | 0.42 | 0.60 | 0.63 | 0.73 | 0.74 | 0.65 | 0.53 | 0.49 | 0.69 | 0.70 | 0.48 | 0.58 | 0.80 | 0.80 |
| (35, 40) | 0.51 | 0.41 | 0.63 | 0.87 | 0.85 | 0.76 | 0.65 | 0.77 | 0.59 | 0.65 | 0.69 | 0.74 | 0.67 | 0.54 | 0.82 | 0.70 |
| (30, 35) | 0.47 | 0.72 | 0.79 | 0.70 | 0.70 | 0.69 | 0.63 | 0.61 | 0.74 | 0.66 | 0.67 | 0.57 | 0.51 | 0.65 | 0.51 | 0.67 |
| (25, 30) | 0.74 | 0.84 | 0.92 | 0.83 | 0.69 | 0.70 | 0.69 | 0.75 | 0.71 | 0.61 | 0.60 | 0.67 | 0.56 | 0.63 | 0.74 | 0.45 |
| (20, 25) | 0.79 | 0.70 | 0.61 | 0.72 | 0.73 | 0.76 | 0.70 | 0.80 | 0.63 | 0.69 | 0.62 | 0.43 | 0.53 | 0.76 | 0.59 | 0.94 |
| (15, 20) | 0.82 | 0.68 | 0.68 | 0.88 | 0.83 | 0.86 | 0.68 | 0.86 | 0.64 | 0.66 | 0.63 | 0.73 | 0.68 | 0.94 | 0.81 | 0.86 |
| (10, 15) | 0.73 | 0.66 | 0.90 | 0.89 | 0.55 | 0.89 | 0.85 | 0.65 | 0.38 | 0.38 | 0.84 | 0.88 | 0.79 | 0.85 | 0.81 | 0.90 |
| (5, 10) | 0.65 | 0.81 | 0.78 | 0.72 | 0.71 | 0.77 | 0.75 | 0.43 | 0.42 | 0.52 | 0.90 | 0.94 | 0.84 | 0.80 | 0.82 | 0.75 |
| (0, 5) | 0.38 | 0.62 | 0.57 | 0.65 | 0.66 | 0.70 | 0.26 | 0.32 | 0.41 | 0.80 | 0.73 | 0.85 | 0.80 | 0.67 | 0.66 | 0.73 |
| | (0, 5) | (5, 10) | (10, 15) | (15, 20) | (20, 25) | (25, 30) | (30, 35) | (35, 40) | (40, 45) | (45, 50) | (50, 55) | (55, 60) | (60, 65) | (65, 70) | (70, 75) | (75, 80) |
