## Appendix B3 for "Quantifying social contact patterns in Minnesota during Stay-at-Home social distancing order"

**Appendix B3.** ATUS vs MN Wave 1 Home. In the percentage change matrix, if there was an increase in number of contacts during the SAH order, that value is coded in blue, if there was a decrease that value is in red. Cells outlined in black represent interactions between respondents and contacts of the same age.

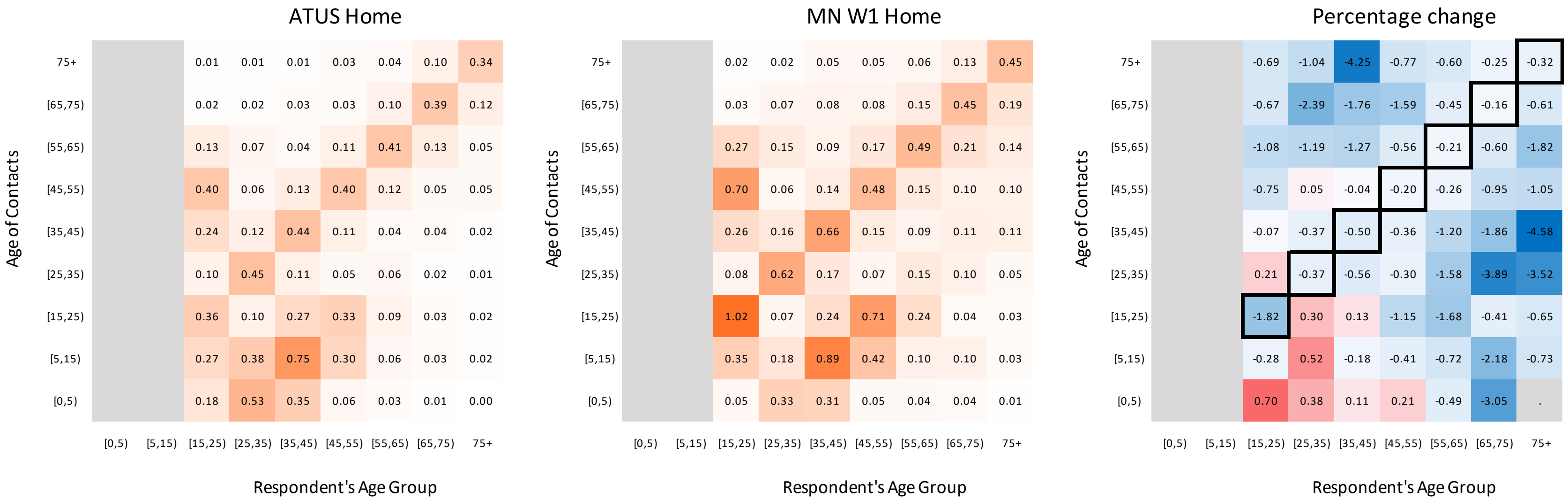
